# Is Covid-19 seroprevalence different in health care workers as per their risk of exposure? A study from a tertiary care hospital in National Capital Region of India

**DOI:** 10.1101/2021.02.10.21251543

**Authors:** Sushila Kataria, Rashmi Phogat, Pooja Sharma, Vikas Deswal, Sazid Alam, Manish Singh, Kuldeep Kumar, Vaibhav Gupta, Padam Singh, Rohit Dutt, Smita Sarma, Renu Saxena, Naresh Trehan

## Abstract

**Background:** SARS-CoV-2 infection has severely ravaged health systems, economic and social progress globally in 2020. Seroprevalence studies can provide relevant information on the target populations for vaccination. They are relevant not only in the community, but also for critical population subgroups such as nursing homes or health care facilities. They will assist in strategizing the vaccination policy especially since there is limited availability of the vaccine and vaccine hesitancy

**Objective:** To evaluate the seroprevalence in Health Care Workers (HCW) at our hospital and to identify parameters which may affect it.

**Methodology:** The Baseline profiling and seroprevalence of severe acute respiratory syndrome coronavirus 2 (SARS-CoV-2) was assessed among 3258 healthcare workers (HCWs) of Medanta-The Medicity, Gurugram, Haryana, India, as a part of an ongoing cohort study.The fully automated LIAISON® SARS-CoV-2 S1/S2 IgG test using the chemiluminescence immunoassay (CLIA) for the quantitative determination of anti-S1 and anti-S2 specific IgG antibodies to SARS-CoV-2 was used to test serum samples collected before the receipt of the vaccine. Seroprevalence was evaluated as per gender, age, association with previous Covid-19 diagnosis, use of supplements, and role in the hospital and type of exposure.

**Results:** Of the 3258 participants tested for IgG serology (S1 and S2 proteins) 46.2% (CI 44.4 – 47.9%) were positive (i.e. had an antibody titre more than 15 Au/ml). Higher seroprevalence was seen in the ‘others’ ie non clinical health care workers (including management, research personnel, pharmacists, technicians, general duty staff, housekeeping, security, food and beverage, and facility maintenance teams) (50.2 Au/ml) than that in clinical HCW (ie doctors and nurses)where it was significantly lower (41.4 Au/ml, p= 0.0001). Also, people with history of Covid-19 were found to have significantly higher antibody levels (p = 0.0001). Amongst the healthcare workers, doctors and nurses had higher relative risk of acquiring Covid-19 infection (RR = 1.21; 95% C.I.: 1.12 - 1.31).

**Conclusion:** Seroprevalence in healthcare workers at our hospital is high at 46.2%. It is higher in non-clinical HCW than in clinical HCW. The risk of acquiring Covid-19 infection was higher in clinical HCW and thus, this subgroup may benefit most from vaccination. History of Covid-19 may provide double the protection, in particular in those who had it recently.

## 1. Introduction

The outbreak of severe acute respiratory syndrome coronavirus 2 (SARS CoV-2), causing the coronavirus disease 2019 (Covid-19), has severely ravaged health systems, economic and social progress globally in 2020. This novel coronavirus spread rapidly to more than 213 countries/territories worldwide and consequently led to a global pandemic (World Health Organization (WHO) since its detection in Hubei province of China in December 2019 [1]. On 30 Jan 2020, WHO declared the novel coronavirus outbreak a public health emergency of international concern (PHEIC), its highest level of alarm [2]. Covid-19 reached the Indian shores of Kerala on Jan 27, 2020 [3]. Worldwide massive responses were put into action, including lockdowns, social distancing measures, quarantine and several drugs, technologies swung into action to minimize morbidity and mortality. Despite all the best efforts of humankind, over 84474195 confirmed cases and 1848704 lives worldwide have been lost to Covid-19 disease [2]. On 23 March 2020, the Government of India ordered a nationwide lockdown. This lockdown and its variants were extended for three more times before the “Unlock” was started on June 8, 2020.The infectious nature of the disease initiated the race to develop and deploy safe and effective vaccine. There are currently more than 50 Covid-19 vaccine candidates in trials [4]. On Dec 11, 2020 the US Food and Drug Administration (FDA) authorized the emergency use of the mRNA vaccine BNT 162b2 to Pfizer and BioNTech, against Covid-19 in individuals 16 years or older in age, this was followed by a national vaccine roll out in the US for high risk persons [5]. On Jan 3, 2021, The Drug Control General of India (DCGI) has approved two Covid-19 vaccines (Oxford-Serum Institute’s Covid-19 Vaccine Covishield and Bharat Biotech’s Covaxin) for restricted emergency use approval (EUA) in India after recommendation by subject expert committee of Central Drugs Standard Control Organization (CDSCO) [6]. On 16 January 2021, India started its national vaccination programme against the <u>SARS-CoV-2</u> which is responsible for the Covid-19 pandemic. The drive prioritises healthcare and frontline workers, and then those over the age of 50 or suffering from certain medical conditions. Several studies have found that the SARS-CoV-2 seroprevalence (the percentage of the population with serum containing antibodies that recognize the virus) has remained below 20% even in the most adversely affected areas globally, such as Spain and Italy [7-9]. The data that is available from India shows a variable seroprevalence in HCW ranging from 2.5% from two different hospitals in Srinagar [10] to 11.94% in a tertiary care center in Kolkata [11].

The variation in rates of seroprevalence being reported from the world and from India, limited availability of the vaccine, and vaccine hesitancy prompted us to evaluate the seroprevalence in HCW at our hospital prior to the national vaccine drive. This paper analyses the baseline seroprevalence findings of a prospective, observational, real world study to evaluate the safety, immunogenicity and effectiveness of the national vaccine roll out.

## 2. Methodology

This study is the part of the baseline evaluation of a larger prospective cohort study evaluating the safety, immunogenicity and effectiveness of the Covid-19 vaccine given as the national vaccine roll out in health care workers. The study was approved by the Institutional Ethics Committee and was conducted in a fifteen hundred bedded tertiary care hospital in the National Capital Region of Delhi, which has treated over ten thousand hospitalized Covid-19 patients. The data was collected between 12 January and 13 February 2021.

The hospital has listed 6962 HCW registered for vaccination in the national vaccine roll out. All health care personnel who consented to participate in the real world cohort for the evaluation of the Covid-19 vaccine were eligible for participation. Further we have excluded those who had participated in any covid prophylactics or drug trials, or had received immunoglobulins and/or convalescent plasma within the three months preceding the planned administration of the vaccine (Jan 16, 2021). Those who were ineligible to receive the vaccine due to history of hypersensitivity reactions, or active Covid-19 disease were also excluded from participation.

The informed consent process followed by a baseline questionnaire was completed by a doctor. The baseline questionnaire collected information on basic socio-demographic characteristics (Age and gender), health information (comorbidities, previous Covid-19 diagnosis, use of any supplements including alternative medicine), and work related information (role in the hospital, the type of exposure at work ie working in a Covid-19 ward, radiology or emergency etc). 3 ml of blood was then collected in Serum Separated Tube (SST, Yellow Top Vacutainer) aseptically by venepuncture. Each sample and form were barcoded, and linked. The samples were stored at centrifuged at 3000 to 3500 RPM for 10 minutes and the separated serum was kept at 2°-8°C until further testing within a week.

### 2.1. Laboratory Methods

The serum was tested for the quantitative determination of anti-S1 and anti-S2 specific IgG antibodies to SARS-CoV-2 in the fully automated LIAISON® SARS-CoV-2 S1/S2 IgG by Chemiluminescence immunoassay (CLIA) technology. The analyzer automatically calculated SARS-CoV-2 S1/S2 IgG antibody concentrations expressed as arbitrary units (AU/mL) and graded the results. The assay detection range is from 3.8 to 400 AU/mL. SARS CoV-2 S1/S2 IgG. < 15 AU/mL were reported negative. Test results >=15.0 AU/mL were graded positive.

### 2.2. Quality Assurance

The fully automated LIAISON® has been calibrated and validated according to the laboratory SOP, ie 3 samples (high, medium and low value) were run 5 times a day for 5 consecutive days. Their mean, SD and CV% were calculated, which were within normal limits. One positive and one negative kit controls were also processed daily for internal quality control. The instrument was calibrated once daily before processing the samples and also for every new lot kit used. Inter-Lab Comparison (ILC) and Repeat - split testing was conducted as per standard laboratory quality assurance protocol.

### 2.3. Data Analysis

For data capture each participant was assigned a unique study code, which facilitated linking of participant data with barcode of serum sample. The data were entered into e-HIS (electronic health information system) template which is protected and exported into Excel sheets. Quality assurance (QA) of the data was taken care of by an independent QA coordinator. Initial analysis was done on the characteristics of the study participants according to role in the hospital, nature of exposure, demographics (age and gender), habits (smoking & alcohol), history of Covid-19 disease, supplements in last 2 months and comorbidities. Detailed analysis of IgG level was attempted in terms of median (IQR) and seroprevalence rate with 95% confidence interval. Further seroprevalence was estimated according to participants role in the hospital, nature of exposure, demographic (age and gender) and history of Covid-19 disease with severity of disease and duration. Percentage at risk has been calculated as compliment of seroprevalence. Thereafter relative risk with respect to reference category (highest protection) has been calculated for participants role in the hospital, nature of exposure, demographic (age and gender) and history of Covid-19 disease with duration. The results have been presented with 95% confidence interval and p values. Mann Whitney Test was used to test the IgG level between participants with and without h/o Covid-19 infection. All analysis was done using SPSS software, version 24.0. P-value < 0.05 has been considered statistically significant.

## 3. Results and Discussion

### 3.1. Results

Out of the total 6962 eligible for vaccination, 3258 (46.8%) agreed to participate in the study (Table 1).

**Table 1:** Representation of participation of hospital staff in the study.

| **Role in the hospital** | **Number at Medanta**<br>**the Medicity** | **Number**<br>**Participated** | **Percent**<br>**(%)** |
| --- | --- | --- | --- |
| **Doctor (Clinical HCW)** | 860 | 342 | 39.8% |
| **Nurse (Clinical HCW)** | 2444 | 1148 | 47.0% |
| **Management (non-clinical HCW)** | 952 | 307 | 32.2% |
| **General duty, Housekeeping, Security,**<br>**Food and Beverage, and facility**<br>**maintenance teams (non-clinical**<br>**HCW)** | 2030 | 1081 | 53.3% |
| **Research personnel and Paramedics**<br>**(non-clinical HCW)** | 676 | 380 | 56.2% |
| **Total** | **6962** | **3258** | **46.8%** |

Of the 3258 participants, 1490 (45.7%) were clinical HCW (doctors and nurses) and 1768 (54.3%) were non-clinical HCW. Research personnel, paramedics, pharmacists, technicians, general duty, housekeeping, security, food and beverage, and facility maintenance teams were included as ‘non-clinical HCW’ for the purpose of this study. 56.4% of the participants were male. 98.5% of the participants were ≤ 60 years old. 2658(78.8%) had a low risk exposure and the remaining were in high risk exposure category as per Ministry of Health and Family Welfare (MoHFW) guidelines [12]. 473 (14.5%) reported a previous history of Covid-19 disease, of which 97.2% reported mild disease (requiring care at covid care centre or home based care 8 (1.7%) had moderate disease, managed in dedicated covid health centres Five people (1.1%) had severe disease with severe pneumonia or ARDS [13]. 328(10.1%) participants were taking supplements in the last three months and these included Vitamin C, Vitamin D, and some Ayurvedic formulations. Only 253 participants reported any comorbidities and the most common of these was Hypertension (89, 2.7%) (Table 2).

**Table 2:**
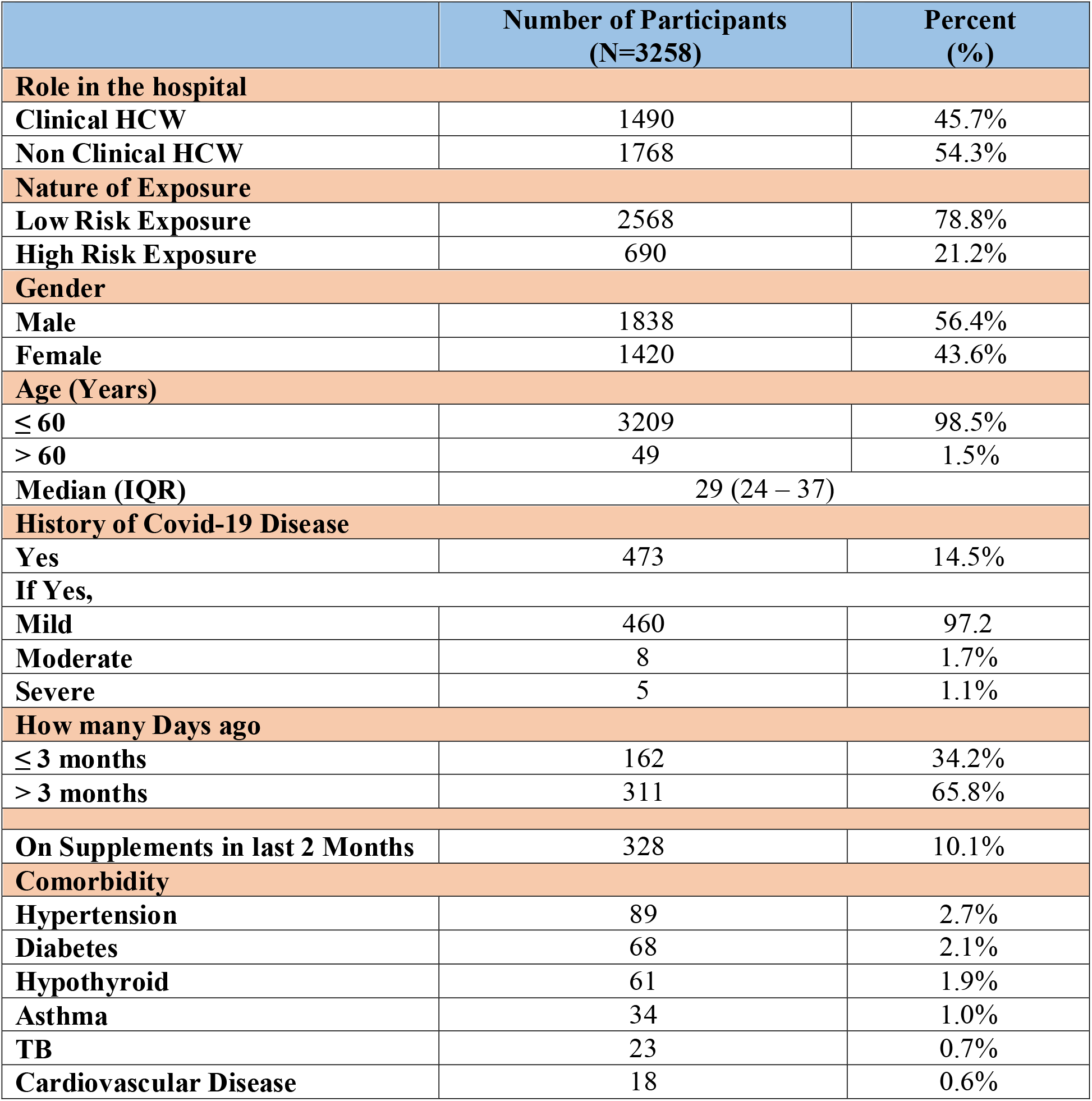
Baseline characteristics of the Study Population (N=3258)

Of the 3258 participants tested for IgG serology (S1 and S2 proteins), 1504 (46.2%) were positive (i.e. had an antibody titre more than/equal to 15 AU/ml) (Table 3 &Figure 1).

**Table 3:** Seroprevalence by IgG>=15 AU /ml (S1 and S2 proteins) n/N = 1504/3258.

| Parameter | Value |
| --- | --- |
| **Covid -19 seroprevalence (≥15 AU/ml)**<br>**n (%) with 95% C. I.** | 46.2% (44.4 – 47.9)% |
| **Value of IgG antibody in those with positive results**<br>**Median (IQR)** | 42.0 (26.9 – 73.2) |

**Figure 1:**
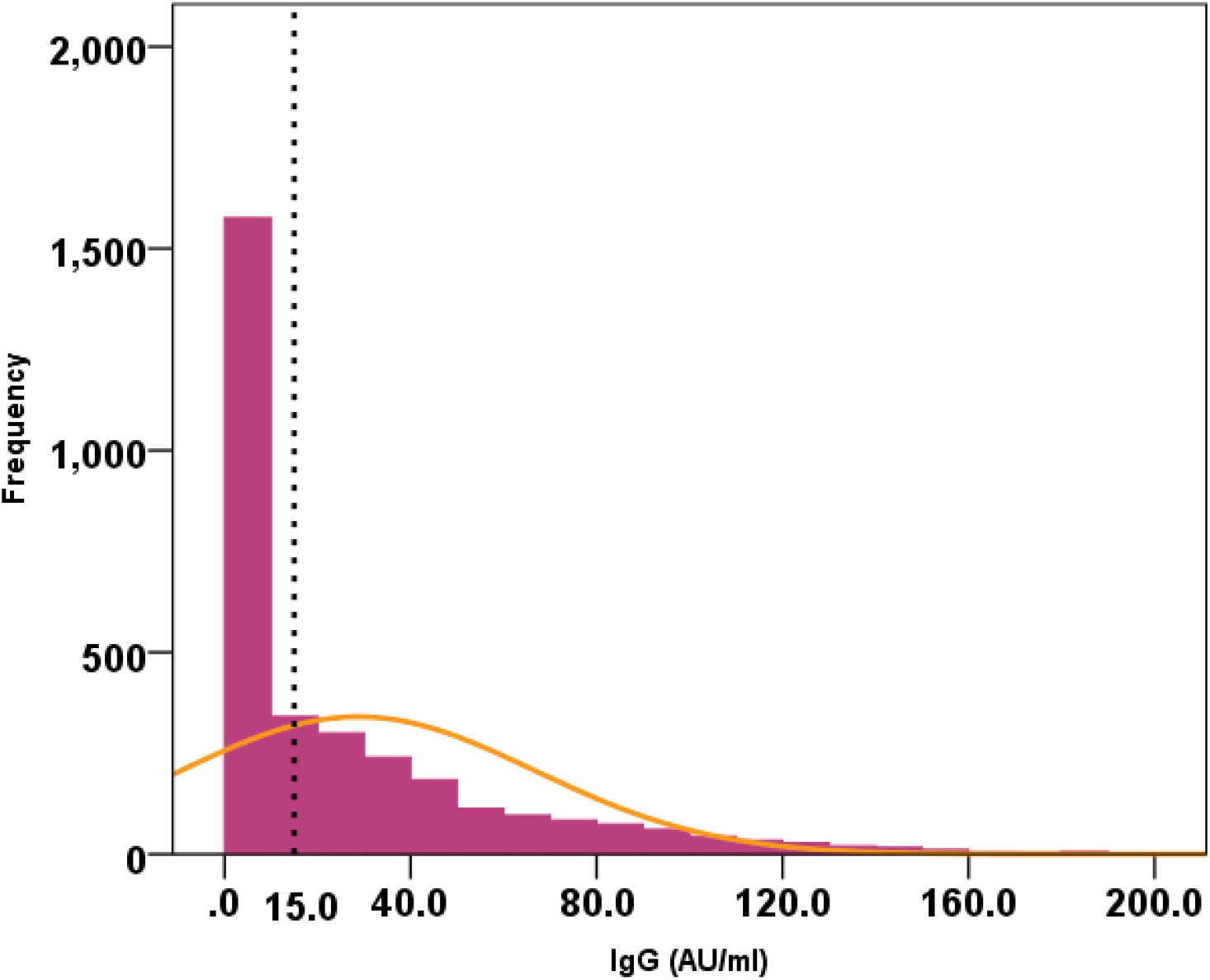
Distribution of the IgG antibody levels in the study (N=3258)

People with a past history of Covid-19 disease were found to have significantly higher antibody levels as compared to those without history of Covid-19 (p = 0.0001) (Figure 2).

**Figure 2:**
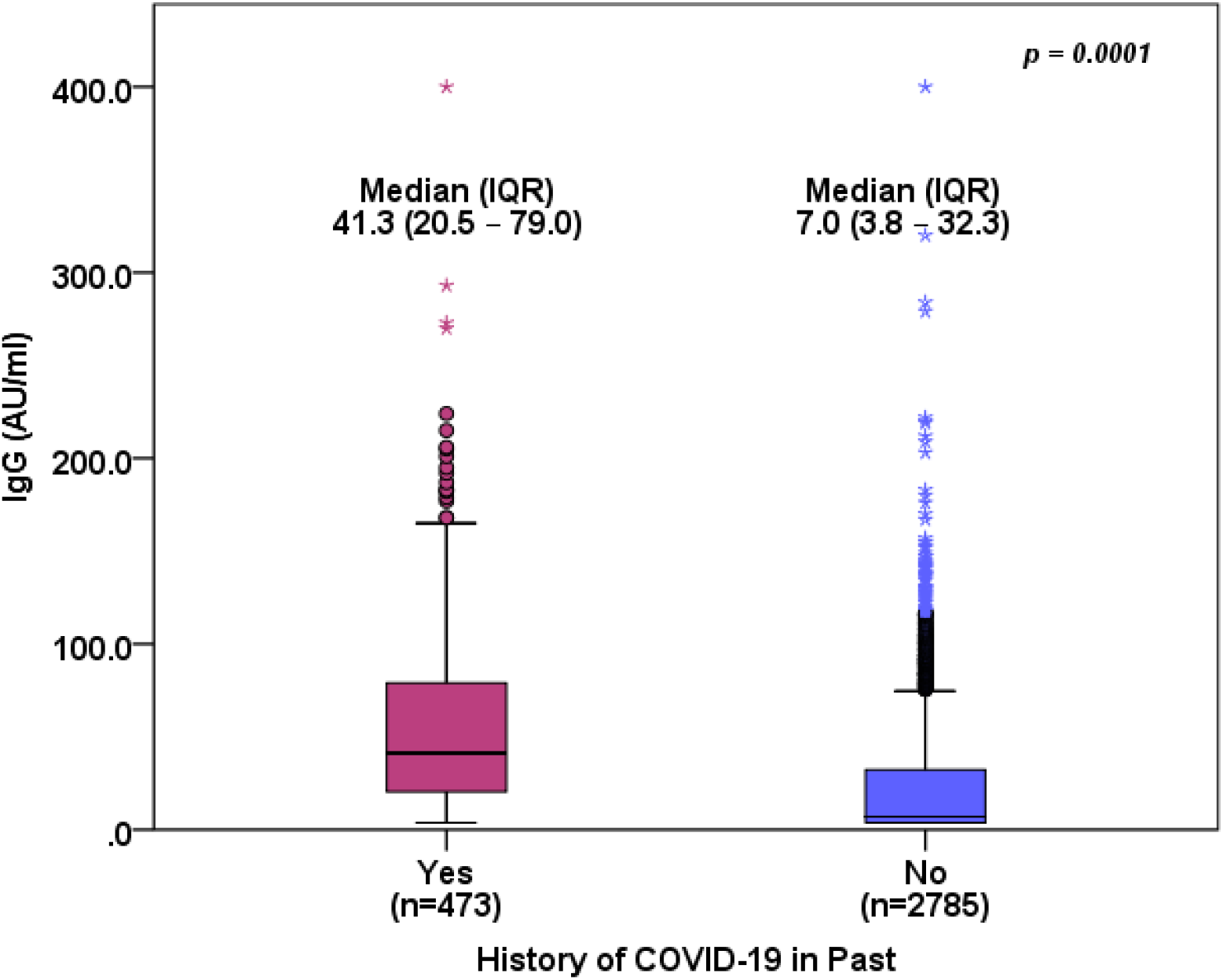
IgG levels in participants with recent history of Covid-19 disease (<3 months)

Seroprevalence was significantly lower in clinical HCW (41.4%) as compared to non-clinical HCW workers (50.2%) (p = 0.0001). There was no significant difference observed in seroprevalence in both low (46.7%) and high (44.3%) risk health care workers (p = 0.262) as well as male (47.1%) and female (44.9%) (p = 0.212). Seroprevalence was significantly lower in age > 60 years (30.6%) as compared to age ≤ 60 years (46.4%) (p = 0.023). Participants with history of Covid-19 were found to have significantly higher seroprevalence as compared to those without history of Covid-19 (81.2% vs. 40.2%; p = 0.0001). Seroprevalence was not significantly different in all the three categories of severity of Covid-19 disease. Seroprevalence was significantly lower in participants who had Covid-19 history before 3 months (75.9%) as compared to those who had Covid-19 history with less than 3 months (91.3%) from the date of survey (p = 0.0001). age > 60 years (30.6%) as compared to age ≤ 60 years (46.4%) (p = 0.023) (Table 4).

**Table 4:** Covid-19 seroprevalence by baseline characteristics (N=3258)

|  | n | Covid-19<br>Seroprevalence<br>(IgG >= 15.0 AU/ml)<br>n (%) | 95% C. I. | p-value |
| --- | --- | --- | --- | --- |
| Role in the hospital |  |  |  |  |
| Clinical HCW | 1490 | 617 (41.4%) | 38.9% - 44.0% | 0.0001* |
| Non Clinical HCW | 1768 | 887 (50.2%) | 47.8% - 52.5% |  |
| Nature of Exposure |  |  |  |  |
| Low Risk Exposure | 2568 | 1198 (46.7%) | 44.7% - 48.6% | 0.262 |
| High Risk Exposure | 690 | 306 (44.3%) | 40.6% - 48.1% |  |
| Gender |  |  |  |  |
| Male | 1838 | 866 (47.1%) | 44.8% - 49.4% | 0.212 |
| Female | 1420 | 638 (44.9%) | 42.3% - 47.6% |  |
| Age Group |  |  |  |  |
| ≤ 60 | 3209 | 1489 (46.4%) | 44.7% – 48.1% | 0.023* |
| > 60 | 49 | 15 (30.6%) | 18.2% - 45.4% |  |
| History of Covid-19 Disease |  |  |  |  |
| No | 2785 | 1120 (40.2%) | 38.4% - 42.1% | 0.0001* |
| Yes | 473 | 384 (81.2%) | 77.4% - 84.6% |  |
| Severity of Covid-19 |  |  |  |  |
| Mild | 460 | 371 (80.6%) | 76.7% - 84.2% | - |
| Moderate | 8 | 8 (100.0%) | 63.1% - 100.0% | 0.166 |
| Severe | 5 | 5 (100.0%) | 47.8% - 100.0% | 0.273 |
| History of Covid-19 Disease (Duration) N=473 |  |  |  |  |
| ≤ 3 months | 162 | 148 (91.3%) | 85.9% – 95.2% | 0.0001* |
| > 3 months | 311 | 236 (75.9%) | 70.7% – 80.5% |  |

**C. I. – Confidence Interval; \*p-value < 0.05, statistically significant**

Clinical HCW are at 21% higher risk as compared to Non Clinical HCW in the hospital. Importantly, history of Covid-19 provides double the protection, in particular those who had it recently. Also those in the age group of 60 years or more have 52% higher risk as compared to those under 60 years of age (Table 5).

**Table 5:**
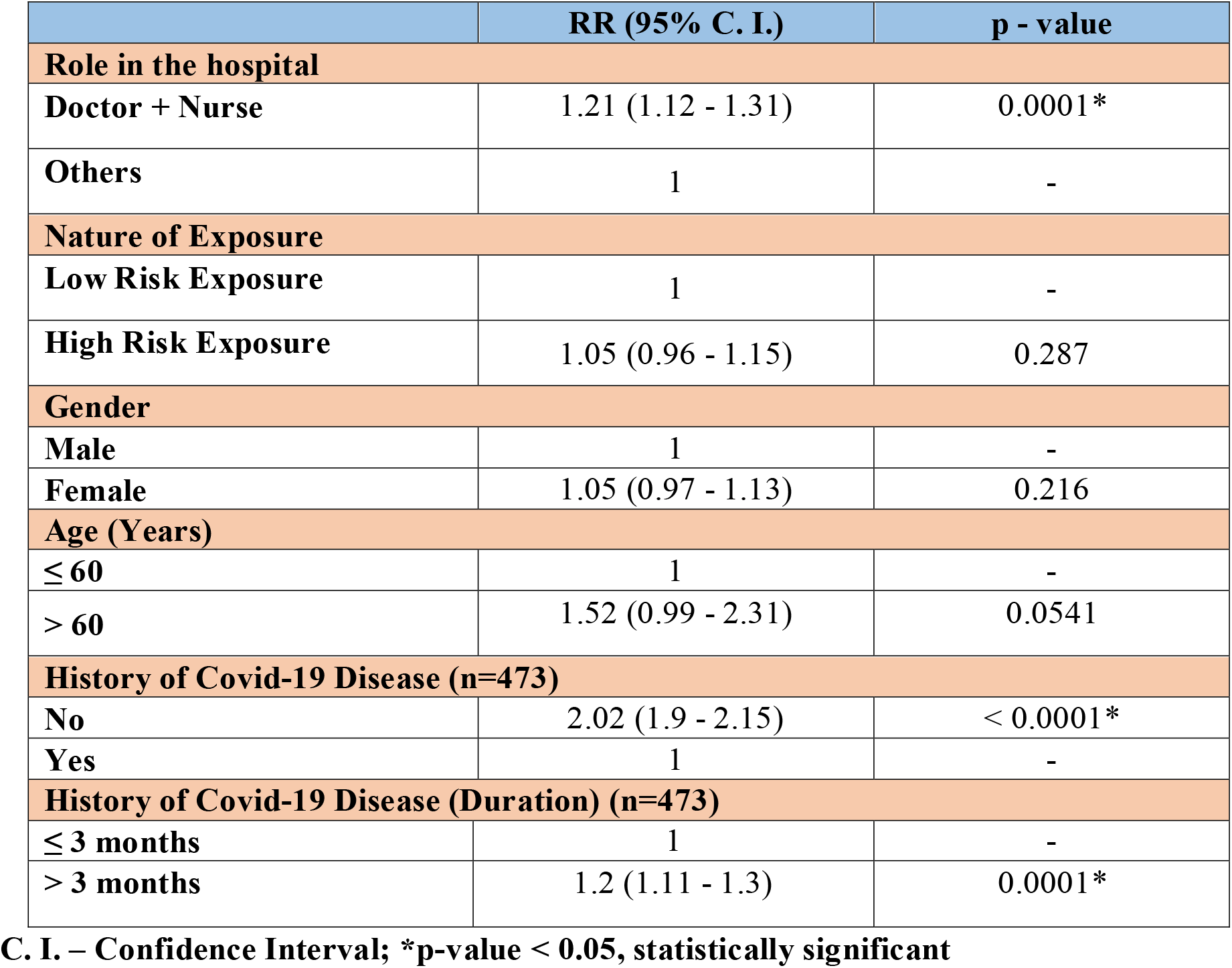
Relative risk of Covid-19 seroprevalence (N=3258)

|  | **RR (95% C. I.)** | **p - value** |
| --- | --- | --- |
| **Role in the hospital** |  |  |
| **Doctor + Nurse** | 1.21 (1.12 - 1.31) | 0.0001* |
| **Others** | 1 | - |
| **Nature of Exposure** |  |  |
| **Low Risk Exposure** | 1 | - |
| **High Risk Exposure** | 1.05 (0.96 - 1.15) | 0.287 |
| **Gender** |  |  |
| **Male** | 1 | - |
| **Female** | 1.05 (0.97 - 1.13) | 0.216 |
| **Age (Years)** |  |  |
| **≤ 60** | 1 | - |
| **> 60** | 1.52 (0.99 - 2.31) | 0.0541 |
| **History of Covid-19 Disease (n=473)** |  |  |
| **No** | 2.02 (1.9 - 2.15) | < 0.0001* |
| **Yes** | 1 | - |
| **History of Covid-19 Disease (Duration) (n=473)** |  |  |
| **≤ 3 months** | 1 | - |
| **> 3 months** | 1.2 (1.11 - 1.3) | 0.0001* |

**C. I. – Confidence Interval; \*p-value < 0.05, statistically significant**

### 3.2. Discussion

The unifying hope for ending the global Covid-19 pandemic is the development of adequate population-level herd immunity to halt the continuing cycles of infection and disease. Although no data exist to define the exact threshold necessary to achieve herd immunity against Covid-19, modelling and extrapolation from similar diseases suggest that more than 60%, and perhaps up to 80%, of the population may need immunity for the viral replication rate to drop below 1, enabling a modest level of disease control [14]. Such immunity may be achieved via recovery of many individuals from widespread infection, or preferably via the availability of safe and effective vaccines. Smallpox remains the only disease in human history to have been eradicated, an achievement of vaccination, not natural immunity [15]. Indeed, there are reasons to be optimistic that prior exposure to the virus does lead to protective immunity. Nearly a year into the Covid-19 pandemic, there have been more than 30 million confirmed infections, but extremely few documented cases of reinfection with SARS-CoV-2 throughout the world [16].

Widespread availability of commercial assays that detect anti–severe acute respiratory syndrome coronavirus 2 (SARS-CoV-2) antibodies has enabled researchers to examine naturally acquired immunity to coronavirus disease 2019 (Covid-19) at the population level. Several studies have found that the SARS-CoV-2 seroprevalence (the percentage of the population with serum containing antibodies that recognize the virus) has remained below 20% even in the most adversely affected areas globally, such as Spain and Italy [7-9]. Bajema *et al* contributed new information on the shifting nature of SARS-CoV-2 seroprevalence in the US [17]. The decline over time of the seroprevalence of antibodies to SARS-CoV-2 in the study by Bajema *et al* is predictable, as for all infectious diseases, the waning of antibody titres is normal and does not necessarily indicate the loss of protective long-term immunity. Immunoglobulin G titres rise during the weeks following infection as active plasma cells secrete antibody into systemic circulation. Those titres then wane as the plasma cells actively secreting the antibodies senesce, whereas resting memory B and T lymphocytes continue to circulate for years to decades and can mediate long-term immunity to infection even in the face of waning antibody titres [18]. Herd immunity is an age-old concept, which is an indirect protection conferred by immune individuals to the susceptible ones in a given population against a specific pathogenic infestation. Herd immunity protects by limiting the spread of the disease [19]. The basic reproductive number (*R*_0_) determines the minimum percentage (*Y*) of the population required to be immune to achieve the herd immunity for the entire population.

As described before, *R*_0_ = 2-3 as per recent reports [17]

If *R*_0_ = 2, then *Y* = [(2 - 1)/2] × 100 = 50%.

Similarly, when *R*_0_ = 3, then *Y* = [(3 - 1)/3 × 100 = 66.66%

Therefore, for *R*_0_ = 2-3, nearly 50% to 66.66%* (threshold) of the population is required to be immune against Covid-19 for the protection of susceptible individuals in a given population through herd immunity. It is unclear how many people have contracted the causative coronavirus (SARS-CoV-2) unknowingly. Signorelli *et al* report seroprevalence in the most effected province of Northern Italy (Bergamo) and its effect on the second wave [20]. The first wave impacted this region particularly violently (1332.9 per 100,000 population). The serosurveillance of 42% Covid-19 antibody positivity was the highest level recorded so far in European seroprevalence studies. It was noted that the second wave impacted Bergamo lesser (257.6 per 100,000 population) than the neighbouring provinces. This may indicate the evolution of an epidemiological picture of herd immunity [21]. Seroprevalence studies help to evaluate the extension of epidemics. Seroprevalence is also an excellent evaluation of the prevention measures of the health care staff [22].

The prevalence and distribution of antibodies to SARS-CoV-2 in a healthy adult populations of various countries such as the Netherlands (2.7%) [23], Turkey (2.7%) [24], Spain (11.2%) [25], a public hospital in New York (27%) [26]. Studies in health care workers showed an equivalently wide spread 14.5% in the Scottish study [27], 1.2% (Japan) [28] 3.66% in Rome [29]. Indian data from two metropolitan studies (Srinagar and Kolkata) show a variation in the prevalence in health care workers ranging from 0.6% to 11.94% [10-11]. Seroprevalence in our study was high at 46.2% (CI 44.4 – 47.9%), this might be due to the changing profile of the seroprevalence in the community from 24.71 % in October 2020 [30] to more than 50% as per Delhi government fifth serosurvey conducted in January, 2021 or due to higher risk of exposure to Covid-19 patients due to the nature of the hospital (focused Covid care hospital). Seroprevalence was significantly lower in clinical HCW (41.4%) as compared to non-clinical HCW workers (50.2%) (p = 0.0001). This may be due to awareness, vigilance and proper use of PPE, other preventive methods in this group of HCW [31]. Seroprevalence was significantly lower in age > 60 years (30.6%) as compared to age ≤ 60 years (46.4%) (p = 0.023) and this has also been reflected in other studies as with advancing age immune response is hampered [32]. Participants with h/o Covid-19 were found to have significantly higher seroprevalence as compared to those without h/o of Covid-19 (81.2% vs. 40.2%; p = 0.0001). Importantly, history of Covid-19 provides double the protection, in particular those who had it recently. Also those in the age group of 60 years or more have 52% higher risk as compared to those under 60 years of age.

## 4.0. Conclusion

Seroprevalence in healthcare workers at our hospital is high at 46.2%. It is higher in non-clinical HCW than in clinical HCW and the risk of acquiring Covid-19 infection was higher in clinical HCW and we feel this subgroup would benefit most from vaccination. History of Covid-19 provides double the protection, in particular those who had it recently.

## Data Availability

All data is available in the hospital

## Acknowledgements

The investigators appreciate the support of Mr Manbir Singh, Senior Lab Technician, Microbiology; Mr Abhinav Yadav, Clinical Research Lab Technician, the Clinical Research team consisting of Mr Dheeraj Sharma, Ms Rita Maity, Mr Rehan Khan, Mr Bajarang, Ms Arunima, Ms Ruchika, Mr Iqbal, Ms Rhea Aggarwal Clinical research and Pharmacy interns from Apeejay Stya University, GD Goenka University, Jamia Hamdard University, MGIMS Sewagram, Mr Rajiv Sikka and the IT team at Medanta, Mr Pankaj Sahni (CEO) and all staff of the Medanta-The Medicity Hospital, Gurugram and all the participants of the research project.

Editorial support in manuscript writing by Mr Radhesh Notiyal

Funding support from an intramural grant of the Medanta Institute of Education and Research

## References

1. World Health Organization. Timeline: WHO’s COVID-19 response. https://www.who.int/emergencies/diseases/novel-coronavirus-2019/interactivetimeline#event-42.

2. World Health Organization. https://www.who.int/emergencies/diseases/novel-coronavirus-2019/situation-reports/ [Accessed 19th January, 2021].

3. Andrews M. A, Areekal B, Rajesh K. R, Krishnan J, Suryakala R, Krishnan B, Muraly C. P, and Santhosh P. V. First confirmed case of COVID-19 infection in India: A case report. Indian J Med Res, 151(5):490–492, 2020. doi:10.4103/ijmr.IJMR_2131_20, 151(5), 490–492.

4. European Medicines Agency. Treatments and vaccines for COVID-19. URL https://www.ema.europa.eu/en/human-regulatory/overview/public-health-threats/coronavirus-disease-covid-19/treatments-vaccines-covid-19. [accessed 7-January-2021].

5. European Medicines Agency decision. URL https://www.ema.europa.eu/en/documents/pip-decision/p/0003/2021-ema-decision-5-january-2021-agreement-paediatric-investigation-plan-granting-deferral-covid-19_en.pdf.

6. Serum Institute of India Obtains Emergency Use Authorisation in India for AstraZeneca’s COVID-19 Vaccine. URL https://www.drugs.com/clinical_trials/serum-institute-india-obtains-emergency-authorisation-india-astrazeneca-s-covid-19-vaccine-19153.html. [accessed 7-January-2021].

7. Percivalle E, Cambiè G, Cassaniti I, Nepita E. V, Maserati R, Ferrari A, Di Martino R, Isernia P, Mojoli F, Bruno R, Tirani M, Cereda D, Nicora C, Lombardo M, and Baldanti F. Prevalence of SARS-CoV-2 specific neutralising antibodies in blood donors from the Lodi Red Zone in Lombardy, Italy, as at 06 April 2020. Euro Surveill, 25(24):2001031, 2020. doi:10.2807/1560-7917.ES.2020.25.24.2001031.

8. Pollán M, Pérez-Gómez B, Pastor-Barriuso R, Oteo J, Hernán MA, Pérez-Olmeda M, Sanmartín JL, Fernández-García A, Cruz I, Fernández de Larrea N, Molina M, Rodríguez-Cabrera F, Martín M, Merino-Amador P, León Paniagua J, Muñoz-Montalvo JF, Blanco F, Yotti R, and ENE-COVID Study Group. Prevalence of SARS-CoV-2 in Spain (ENE-COVID): a nationwide, population-based seroepidemiological study. Lancet, 22;396(10250):535–544, 2020.

9. Havers F. P, Reed C, Lim T, Montgomery J. M, Klena J. D, Hall A. J, Fry A. M, Cannon D. L, Chiang C. F, Gibbons A, Krapiunaya I, Morales-Betoulle M, Roguski K, Rasheed M. A. U, Freeman B, Lester S, Mills L, Carroll D. S, Owen S. M, Johnson J. A, Semenova V, Blackmore C, Blog D, Chai S. J, Dunn A, Hand J, Jain S, Lindquist S, Lynfield R, Pritchard S, Sokol T, Sosa L, Turabelidze G, Watkins S. M, Wiesman J, Williams R. W, Yendell S, Schiffer J, and Thornburg N. J. Seroprevalence of Antibodies to SARS-CoV-2 in 10 Sites in the United States, March 23-May 12, 2020. JAMA Intern Med, 2020. doi:10.1001/jamainternmed.2020.4130.

10. Khan M. S, Haq I, Qurieshi M. A, Majid S, Bhat A.A, Obaid M, Qazi T. B, Chowdri N, Sabah I, Kawoosa M. F., Lone A. A, Nabi S, Sumji I. A, and Kousar R. SARS-CoV-2 seroprevalence in healthcare workers of dedicated-COVID hospitals and non– COVID hospitals of District Srinagar, Kashmir. medRxiv 10.23.20218164, 2020. doi:https://doi.org/10.1101/2020.10.23.20218164.

11. Goenka M, Afzalpurkar S, Goenka U, Das S. S, Mukherjee M, Jajodia S, Shah B. B, Patil V. U, Rodge G, Khan U, and Bandyopadhyay S. Seroprevalence of COVID-19 Amongst Health Care Workers in a Tertiary Care Hospital of a Metropolitan City from India. J Assoc Physicians India. 68(11):14–19, 2020. PMID: 33187030.

12. https://www.mohfw.gov.in/pdf/AdvisoryformanagingHealthcareworkersworkinginCOVIDandNonCOVIDareasofthehospital.pdf.

13. https://www.mohfw.gov.in/pdf/UpdatedClinicalManagementProtocolforCOVID19dated03072020.pdf.

14. Fontanet A, and Cauchemez S. COVID-19 herd immunity: where are we? Nat Rev Immunol. 20(10):583–584, 2020. doi:10.1038/s41577-020-00451-5.

15. Spellberg B, Nielsen T. B, and Casadevall A. Antibodies, Immunity, and COVID-19. JAMA Intern Med, 2020. doi:10.1001/jamainternmed.2020.7986.

16. Tillett R. L, Sevinsky J. R, Hartley P. D, Kerwin H, Crawford N, Gorzalski A, Laverdure C, Verma S. C, Rossetto C. C, Jackson D, Farrell M. J, Hooser S. V, and Pandori M. Genomic evidence for reinfection with SARS-CoV-2: a case study. Lancet Infect Dis. 21: 52–58, 2020. PIIS1473-3099(20)30764-7.

17. Bajema K. L, Wiegand R. E, Cuffe K, Patel S. V, Iachan R, Lim T, Lee A, Moyse D, Havers F. P, Harding L, Fry A. M, Hall A. J, Martin K, Biel M, Deng Y, Meyer W. A 3rd, Mathur M, Kyle T, Gundlapalli A. V, Thornburg N. J, Petersen L. R, and Edens C. Estimated SARS-CoV-2 Seroprevalence in the US as of September 2020. JAMA Intern Med. e207976, 2020. doi:10.1001/jamainternmed.2020.7976.

18. Syal K. COVID-19: Herd immunity and convalescent plasma transfer therapy. J Med Virol. 92:1380–1382, 2020. https://doi.org/10.1002/jmv.25870.

19. Liu Y, Gayle A. A, Wilder-Smith A, and Rocklöv J. The reproductive number of COVID-19 is higher compared to SARS coronavirus. J Travel Med. 27(2):taaa021, 2020. doi:10.1093/jtm/taaa021.

20. Signorelli C, Zucchi A, Tersalvi CA, Ciampichini R, Beato E, Balzarini F, Odone A, and Middleton J. High seroprevalence of SARS_COV-2 in Bergamo: evidence for herd immunity or reason to be cautious? Int J Public Health. 65(9):1815–1817, 2020. doi:10.1007/s00038-020-01524-x.

21. Bubar KM, Reinholt K, Kissler SM, Lipsitch M, Cobey S, Grad YH, and Larremore DB. Model-informed COVID-19 vaccine prioritization strategies by age and serostatus. Science. eabe6959, 2021. doi:10.1126/science.abe6959.

22. Raoult D. How useful is serology for COVID-19? Int J Infect Dis. 102:170–171, 2021. doi:10.1016/j.ijid.2020.10.058.

23. Slot E, Hogema BM, Reusken Cbem, Reimerink JH, Molier M, Karregat JHM, IJlst J, Novotný Vmj, van Lier Raw, Zaaijer HL. Low SARS-CoV-2 seroprevalence in blood donors in the early COVID-19 epidemic in the Netherlands. Nat Commun. 11(1): 5744, 2020. doi:10.1038/s41467-020-19481-7.

24. Gizem A, Ahmet M, Zeki A, Ozge T, Nihat B. A, Arzu I, Mehtap A, Ridvan K, Mustafa G, Batuhan Y, Hasan T, Adil M, Mehmet D, Filiz B, Nurhan S, Gizem D. D, and Levent D. Seroprevalence of Coronavirus Disease 2019 (COVID-19) Among Health Care Workers from Three Pandemic Hospitals of Turkey. medRxiv. 20178095, 2020. doi:https://doi.org/10.1101/2020.08.19.20178095.

25. Garcia-Basteiro A. L, Moncunill G, Tortajada M, Vidal M, Guinovart C, Jiménez A, Santano R, Sanz S, Méndez S, Llupià A, Aguilar R, Alonso S, Barrios D, Carolis C, Cisteró P, Chóliz E, Cruz A, Fochs S, Jairoce C, Hecht J, Lamoglia M, Martínez M. J, Mitchell R. A, Ortega N, Pey N, Puyol L, Ribes M, Rosell N, Sotomayor P, Torres S, Williams S, Barroso S, Vilella A, Muñoz J, Trilla A, Varela P, Mayor A, and Dobaño C. Seroprevalence of antibodies against SARS-CoV-2 among health care workers in a large Spanish reference hospital. Nat Commun. 11(1):3500, 2020. doi:10.1038/s41467-020-17318-x.

26. Venugopal U, Jilani N, Rabah S, Shariff M. A, Jawed M, Mendez Batres A, Abubacker M, Menon S, Pillai A, Shabarek N, Kasubhai M, Dimitrov V, and Menon V. SARS-CoV-2 seroprevalence among health care workers in a New York City hospital: A cross-sectional analysis during the COVID-19 pandemic. Int J Infect Dis. 102:63–69, 2021. doi:10.1016/j.ijid.2020.10.036.

27. Abo-Leyah H, Gallant S, Cassidy D, Giam Y. H, Killick J, Marshall B, Hay G, Pembridge T, Strachan R, Gallant N, Parcell B. J, George J, Furrie E, and Chalmers J. D. Seroprevalence of SARS-COV-2 Antibodies in Scottish Healthcare Workers. medRxiv. 20205641, 2020. doi:https://doi.org/10.1101/2020.10.02.20205641.

28. Yoshihara T, Ito K, Zaitsu M, Chung E, Aoyagi I, Kaji Y, Tsuru T, Yonemura T, Yamaguchi K, Nakayama S, Tanaka Y, Yurino N, Koyanagi H, Matsuki S, Urae R, and Irie S. SARS-CoV-2 seroprevalence among healthcare workers in general hospitals and clinics in Japan. medRxiv. 21249922, 2021. doi:https://doi.org/10.1101/2021.01.23.21249922.

29. Vetrugno G, La Milia D. I, D’Ambrosio F, Pumpo M. D, Pastorino R, Boccia S, Ricci R, De-Giorgio F, Cicconi M, Foti F, Pascucci D, Castrini F, Carini E, Cambieri A, D’Alfonso M. E, Capalbo G, Fantoni M, Moscato U, Staiti D, De Simone F. M, Berloco F, Zega M, Cattani P, Posteraro B, Sanguinetti M, and Laurenti P. COVID-19 seroprevalence among healthcare workers of a large COVID Hospital in Rome reveals strengths and limits of two different serological tests. medRxiv. 21249445, 2021. doi:https://doi.org/10.1101/2021.01.08.21249445.

30. Sharma N, Sharma P, Basu S, Saxena S, Chawla R, Kumar Det al. The seroprevalence and trends of SARS-CoV-2 in Delhi, India: A repeated population based seroepidemiological study. medRxiv, 20248123, 2020. doi:https://doi.org/10.1101/2020.12.13.20248123.

31. Liu Min, Cheng Shou-Zhen, Xu Ke-Wei, Yang Yang, Zhu Qing-Tang, Zhang Hui et al. Use of personal protective equipment against coronavirus disease 2019 by healthcare professionals in Wuhan, China: cross sectional study BMJ 2020; 369 :m2195

32. Chen J, Kelley WJ, Goldstein DR. Role of aging and the immune response to respiratory viral infections: potential implications for COVID-19. The Journal of Immunology. 2020 Jul 15;205 (2):313–20.

